# Prevalence and Factors Associated with Mental Illness Symptoms among School Students Post Lockdown of the COVID-19 Pandemic in the United Arab Emirates: A Cross-Sectional National Study

**DOI:** 10.1101/2022.07.20.22277866

**Authors:** Nariman Ali Ghader, Noor Al Mheiri, Asma Fikri, Hira AbdulRazzak, Hassan Saleheen, Basema Saddik, Yousef Aljawarneh, Heyam Dalky, Ammar Al Banna, Shammah Al Memari, Budoor Al Shehhi, Shereena Al Mazrouei, Omniyat Al Hajeri

**Author notes:** Corresponding author: Nariman Ali Ghader (NAG).

## Abstract

Data from the United Arab Emirates about the mental health status of the younger population is limited. The current study aimed to assess the prevalence of anxiety, depression, and risk for PTSD among school students post lockdown of the current COVID-19 pandemic. A sample of 3745 students and their parents across the country answered a web-based survey. Parents provided bio-demographic data and students answered questions from the Mood and Feeling Questionnaire (MFQ-Child Self-report), Screen for Child Anxiety Related Disorders (SCARED-Child Version), and Children’s Revised Impact of Event Scale (CRIES-8). Findings showed that the risk for PTSD was the most prevalent (40.6%), followed by symptoms of anxiety (23.3%), and depression (17.1%). For gender differences, symptoms of the three conditions were higher in female students by 6.9%. Moreover, symptoms of depression and anxiety were found to be higher among late adolescents. Further analysis revealed that having medical problems (β = 2.0, *p* < 0.001) and witnessing the death of a close family member due to COVID-19 (β = 1.7, *p* < 0.001) were positive predictors associated with PTDS, depression, and anxiety. The study concluded that post COVID-19 lockdown, symptoms of anxiety, depression, and risk for PTSD are prevalent among students in the UAE. Researchers recommend the initiation of a national school mental health screening program and the provision of follow-up services for vulnerable students. Another must-have is the integration of a mental health support system in the emergency and disaster preparedness future plans.

## Introduction

Since the COVID-19 pandemic emerged, people and governments have been overwhelmed by the sudden rise in widespread mortality, morbidity, and disruption of everyday routines. Mental health has been considered a pivotal area of concern over the course of this particular period and the World Health Organization (WHO) affirmed that 90% of countries experienced substantial disruptions to basic healthcare services and a notable increase in demand for mental health services. Globally, the prevalence of depression, stress, and anxiety among the general population during the COVID-19 pandemic is estimated to be 33.7%, 29.6%, and 31.9% respectively [1]. The growing threat of contracting the infection, the tragic stories of suffering and death, the imposed restrictions and policies (social isolation, lockdowns, vaccinations, etc…), the mania for information in the media, and the economic consequences have created a worldwide climate of depression and anxiety. Studies have revealed an increased risk of post-traumatic stress disorder (PTSD), sleep disorders, and cognitive underperformance in comparison to the pre-pandemic period [2].

Conventionally, children and young people are at the heart of global development. However, in a press release by UNICEF in October 2021, it was affirmed that the younger generation has been carrying the burden of mental health conditions even prior to COVID-19. Globally, one in seven adolescents aged 10-19 years is estimated to suffer from a mental disorder according to the State of the World’s Children 2021 report. Undeniably, the complexity of this state has been accentuated by the aggressions of the pandemic. Additionally, the report warned that children and young people may experience the impact of COVID-19 on their mental health and well-being for many years to come [3]. Further evidence showed that COVID-19 and its related consequences have had a significant impact on youth and their psychological well-being. Uncertainties associated with the pandemic itself, such as prolonged and widespread parental stressors, school closures, and loss of loved ones have been detrimental to the well-being of adolescents and children [4].

Information about the impact of COVID-19 on the mental health of the United Arab Emirates (UAE) population is limited [5]. In an earlier study about the consequences of the pandemic on the psychological well-being of university students, it was revealed that more than 50% of students reported anxiety levels that ranged from mild to severe with females reporting higher levels of anxiety (OR=2.02, 95% CI, 1.41 to 2.91) [6]. Another study conducted in the UAE explored the psychosocial correlates (mental health history, COVID-19 infection status, demographic variables, and living arrangements) with depression and anxiety in a community of adults. Results showed higher levels of anxiety and depression as compared to the pre-pandemic figures. Furthermore, statistically significant associations were found between depression and anxiety on one hand, and being young, female, having a history of mental health problems, and having oneself or loved ones tested positive for COVID-19 on the other [7].

Indisputably, studies on the mental health of the UAE’s younger population are essential as 17-22% of the nation’s youth suffer from depressive symptoms [8]. Expectedly, the emergence of COVID-19 has exacerbated this state of disturbance. It has been acknowledged that during the pandemic, children in the UAE were facing increased mental health issues associated with stories of calamities, home confinement, and fear of contracting the virus. Furthermore, this global crisis was found to have affected negatively children’s mental, behavioral, physical, sleep, and dietary health in the UAE [9, 10].

This national study, which was carried out by a consortium of government health entities and academia within the UAE, aimed at establishing a robust data reference on the prevalence of mental illness symptoms among the younger population. The findings, which can be attributed to the current pandemic, will serve as scientific evidence on the magnitude and nature of work needed in children and adolescent mental health. Furthermore, this study will guide strategists and policymakers in the development of emergency and disaster preparedness plans and provide a clearer path for accurate future foresight.

## Objective

This study aimed to investigate the prevalence of mental illness symptoms (depression, anxiety, and PTSD) among school students in the UAE post lockdown of the COVID-19 pandemic. It was hypothesized that mental illness symptoms are highly prevalent among school students in the UAE which could be exacerbated by the current outbreak.

## Method

In this exploratory cross-sectional study, all students in the UAE private and government schools from grades 2 to 12, aged between 8 and 18 years, and believed to be capable of reading and understanding adequately written Arabic and/or English texts were invited to take in this study. However, students who were enrolled in vocational, continuing education, or special education programs/ schools were excluded. Equally well, parents of the surveyed students participated in the study by consenting on behalf of their under-age children and answering a series of questions within the same instrument.

### Data collection procedure

Invites for participation were sent out to the public via multiple corporate channels including SMS, press release, and social media platforms. In addition, school-parent communication channels served as a fundamental resource for reaching out to the study participants. Parents and their children were asked to complete an online questionnaire using the UAE government survey platform “mSurvey”. Data was collected after students passed through a period of lockdown, however, the majority (83.3%) were still enrolled in total online school learning. Responses were collected over a period of six months (February – July 2021).

Depending on the students’ reading and comprehension abilities, parents were instructed to help their children read and understand the questions while avoiding the interpretation of answers. In addition, if a household had two or more younger children, parents were advised to help each child complete the questionnaire separately.

### The instrument

The data collection instrument used in this study was composed of three sections. Firstly, a detailed participant information sheet and an informed consent form were mandatory fields to be completed before participants had access to the anonymous questionnaire. Secondly, parents were asked to respond to 26 questions which included family sociodemographic data as well as information on the student’s general health status, experience with COVID-19 infection, school learning, and mental health-related behaviors; the estimated time to complete this part was 15 minutes. Thirdly, students were also requested to answer over an estimated 30 minute-period a series of 8 questions that explored their feelings and their current mental health status. Data on students’ mental health-related behaviors were collated via a collection of standardized scales. MFQ-Child self-report (13-items), SCARED-Child Version (41-items), and CRIES were used to screen children for current depressive symptomatology, anxiety-related disorders and a wide range of traumatic events respectively [11-14].

### Privacy and Confidentiality

Data were collected anonymously and were confidentially stored in the Emirates Health Services database system. Investigators and statisticians who helped in data analysis were the only individuals authorized to access the data and only after signing a non-disclosure agreement form.

### Ethical Considerations

Ethical approvals were acquired from the Emirates Institutional Review Board for COVID-19 Research (Reference No. DOH/CVDC/2020/2471) and the Research Ethics Committee at the Emirates Health Services, Dubai (reference No. MOHAP/DXB-REC/ DDD/No.163 /2020). Moreover, anonymity, confidentiality, and voluntariness were preserved throughout the study. As is conventional in anonymous surveys, consent was in place before starting the survey. Parents represented their underage children in accepting participation in this study. Parents confirmed their consent by answering a mandatory question prior to beginning the survey. Participants were informed they could stop completing the questionnaire or refrain from submitting their responses if they felt uncomfortable at any stage during completion. Participation in the study was voluntary. Participants had the full right to withdraw at any time without the need to justify their actions. There were no apparent risks that could result from participating in this study. However, due to the fact that some students could feel uncomfortable expressing their feelings, a portal for remote counseling services was communicated to all participants.

### Data Analysis

Data were analyzed using IBM SPSS Statistics for Windows version 26 (IBM Corp., Armonk, N.Y., USA). The number of submitted questionnaires was 3762. However, post-data cleaning, the number was reduced to 3745. Mean and standard deviation (SD) were calculated for continuous variables while proportions (%) and frequencies (n) were reported for categorical variables. A multivariate analysis was conducted to examine the associations between mental illness symptoms and related factors by using the Pearson’s product-moment correlation coefficient test. Multiple linear regression analyses were performed to examine the effects of selected demographic and COVID-19 related factors on students’ mental illness symptoms scores. A *P*-value < 0.05 was considered statistically significant in all analyses.

### Findings

The socio-demographic characteristics of participants are shown in Table 1. The majority of parents were married (94.2%), employed (81.4%), and expats (67.8%). Likewise, most respondents (93.5%) had none of their family members been previously diagnosed with a mental health-related problem or behavioral disorder. As for students, over one-third (34.3%) were pre-adolescents (<10 yrs.), 37.5% belonged to the early adolescence age group (10-13 yrs.), 23.7% to middle adolescence (14-17 yrs.), and 1.8% to late adolescence (18 yrs.). For gender, a little over half (50.6%) were male. Most of the students (81.6%) studied in private schools and attended school completely online (82.3%).

**Table 1:**
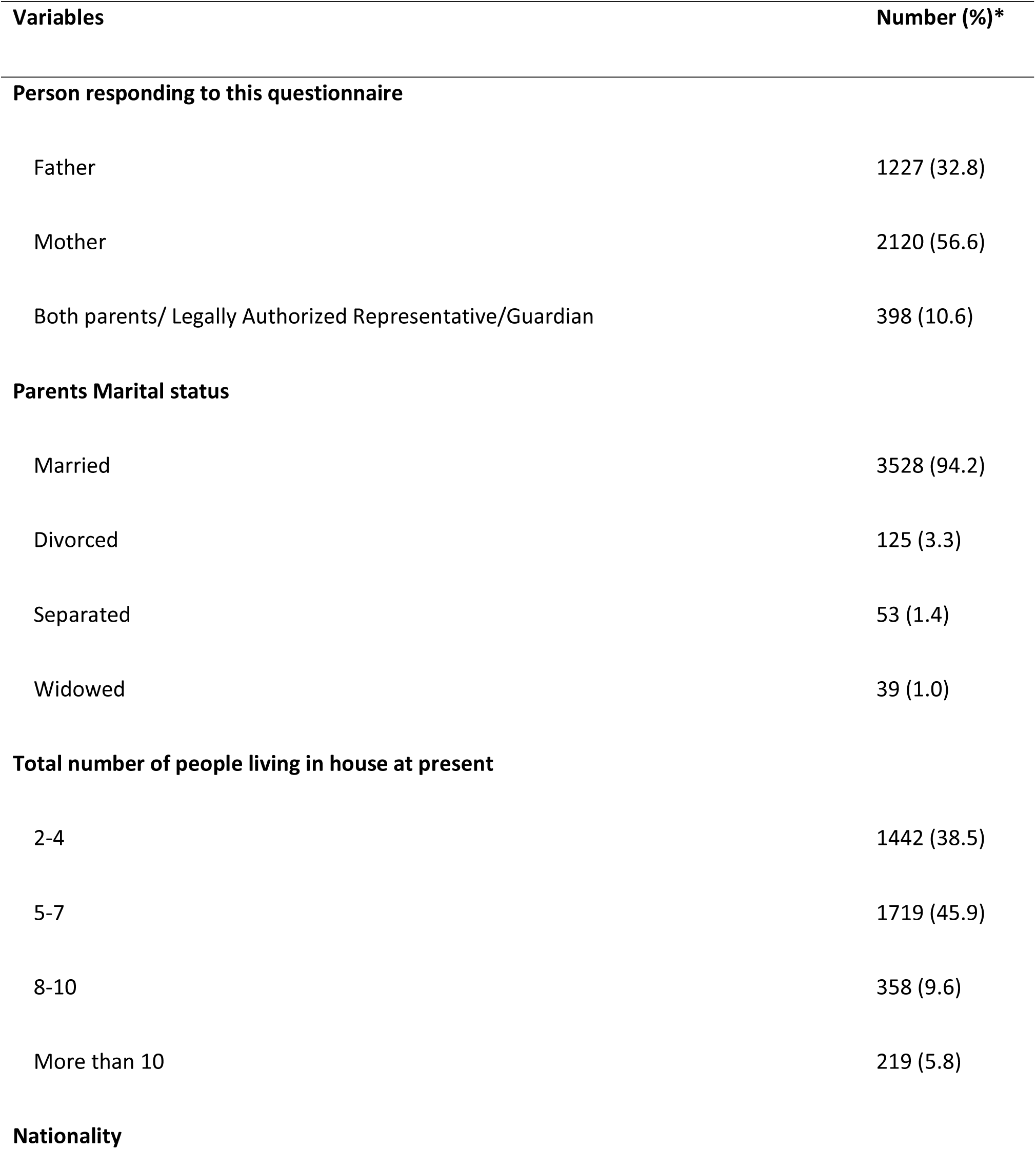
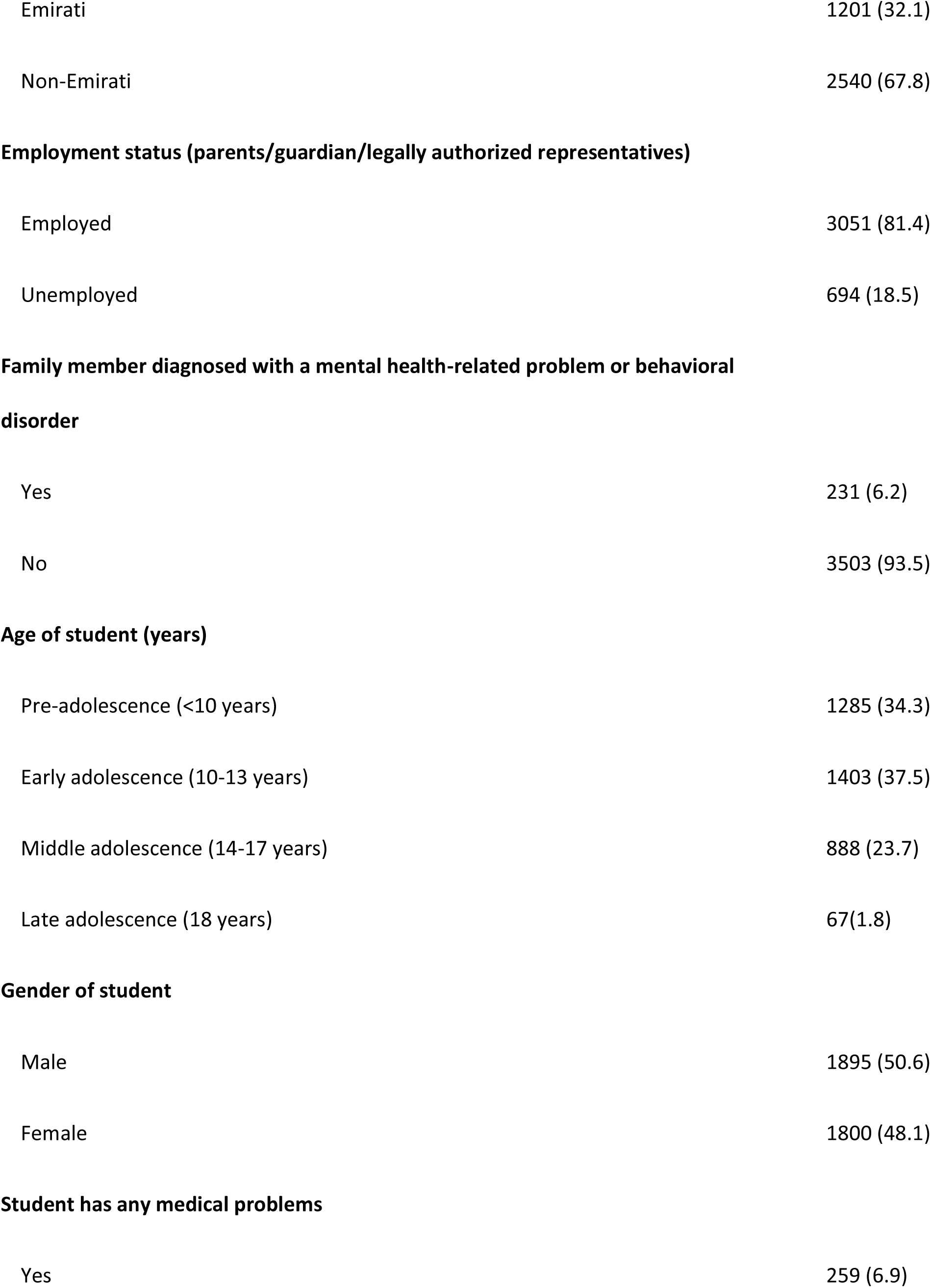
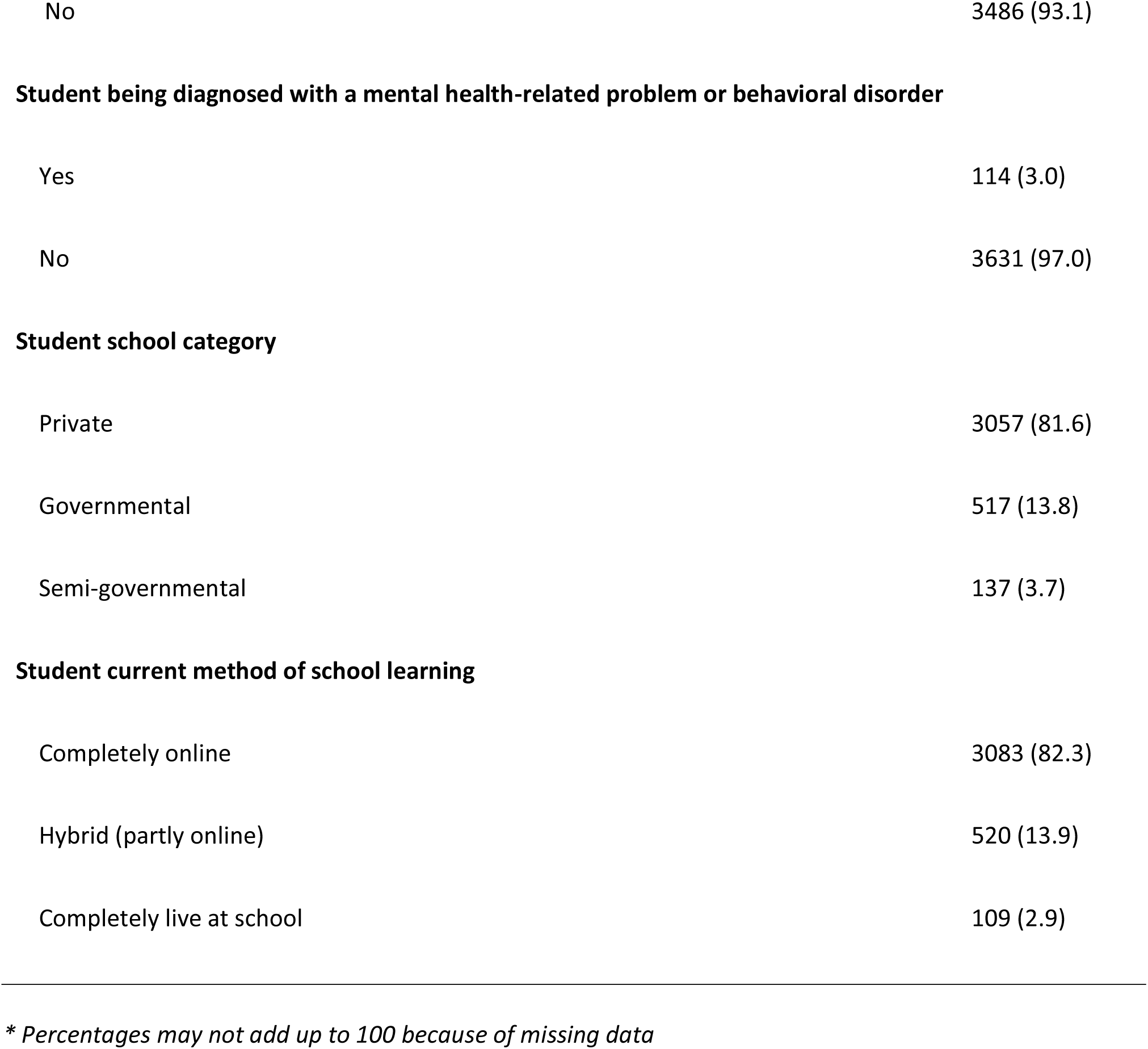
Socio-demographic characteristics of the sample (N=3,745)

| Variables | Number (%)* |
| --- | --- |
| **Person responding to this questionnaire** |  |
| Father | 1227 (32.8) |
| Mother | 2120 (56.6) |
| Both parents/ Legally Authorized Representative/Guardian | 398 (10.6) |
| **Parents Marital status** |  |
| Married | 3528 (94.2) |
| Divorced | 125 (3.3) |
| Separated | 53 (1.4) |
| Widowed | 39 (1.0) |
| **Total number of people living in house at present** |  |
| 2-4 | 1442 (38.5) |
| 5-7 | 1719 (45.9) |
| 8-10 | 358 (9.6) |
| More than 10 | 219 (5.8) |
| **Nationality** |  |
| Emirati | 1201 (32.1) |
| Non-Emirati | 2540 (67.8) |
| **Employment status (parents/guardian/legally authorized representatives)** |  |
| Employed | 3051 (81.4) |
| Unemployed | 694 (18.5) |
| **Family member diagnosed with a mental health-related problem or behavioral disorder** |  |
| Yes | 231 (6.2) |
| No | 3503 (93.5) |
| **Age of student (years)** |  |
| Pre-adolescence (<10 years) | 1285 (34.3) |
| Early adolescence (10-13 years) | 1403 (37.5) |
| Middle adolescence (14-17 years) | 888 (23.7) |
| Late adolescence (18 years) | 67(1.8) |
| **Gender of student** |  |
| Male | 1895 (50.6) |
| Female | 1800 (48.1) |
| **Student has any medical problems** |  |
| Yes | 259 (6.9) |
| No | 3486 (93.1) |
| **Student being diagnosed with a mental health-related problem or behavioral disorder** |  |
| Yes | 114 (3.0) |
| No | 3631 (97.0) |
| **Student school category** |  |
| Private | 3057 (81.6) |
| Governmental | 517 (13.8) |
| Semi-governmental | 137 (3.7) |
| **Student current method of school learning** |  |
| Completely online | 3083 (82.3) |
| Hybrid (partly online) | 520 (13.9) |
| Completely live at school | 109 (2.9) |

*\* Percentages may not add up to 100 because of missing data*

The epidemic-related data of the study sample showed that 14% had a close family member at the time tested positive for coronavirus, followed by 14% who indicated that a close family member was before or at the time sick/hospitalized because of the current coronavirus infection. However, only 5.8% reported that a close family member had died because of the coronavirus. A smaller percentage of students were declared to be testing positive for coronavirus at the time (3%) while 5.4% were sick or hospitalized (Table 2).

**Table 2:**
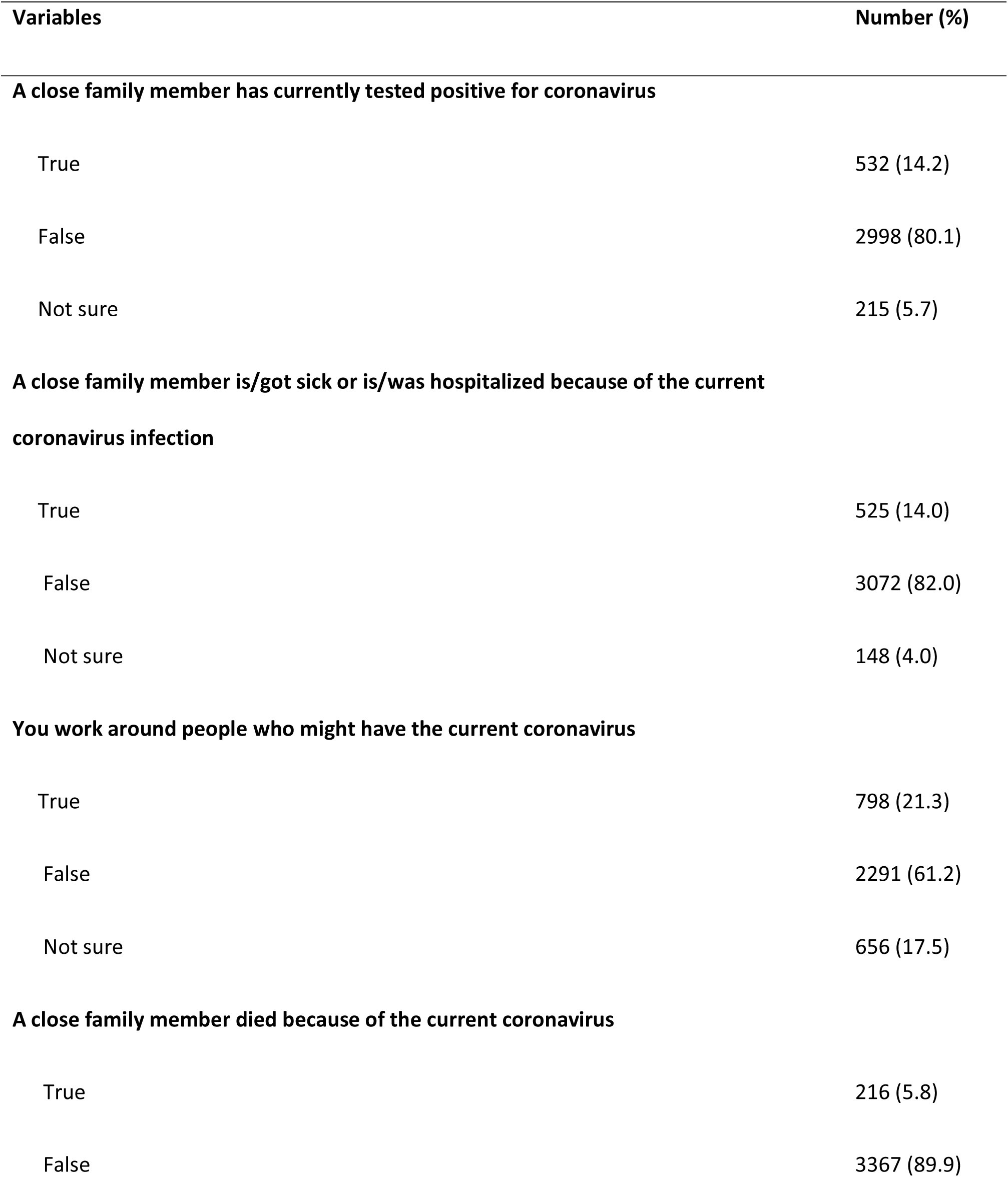
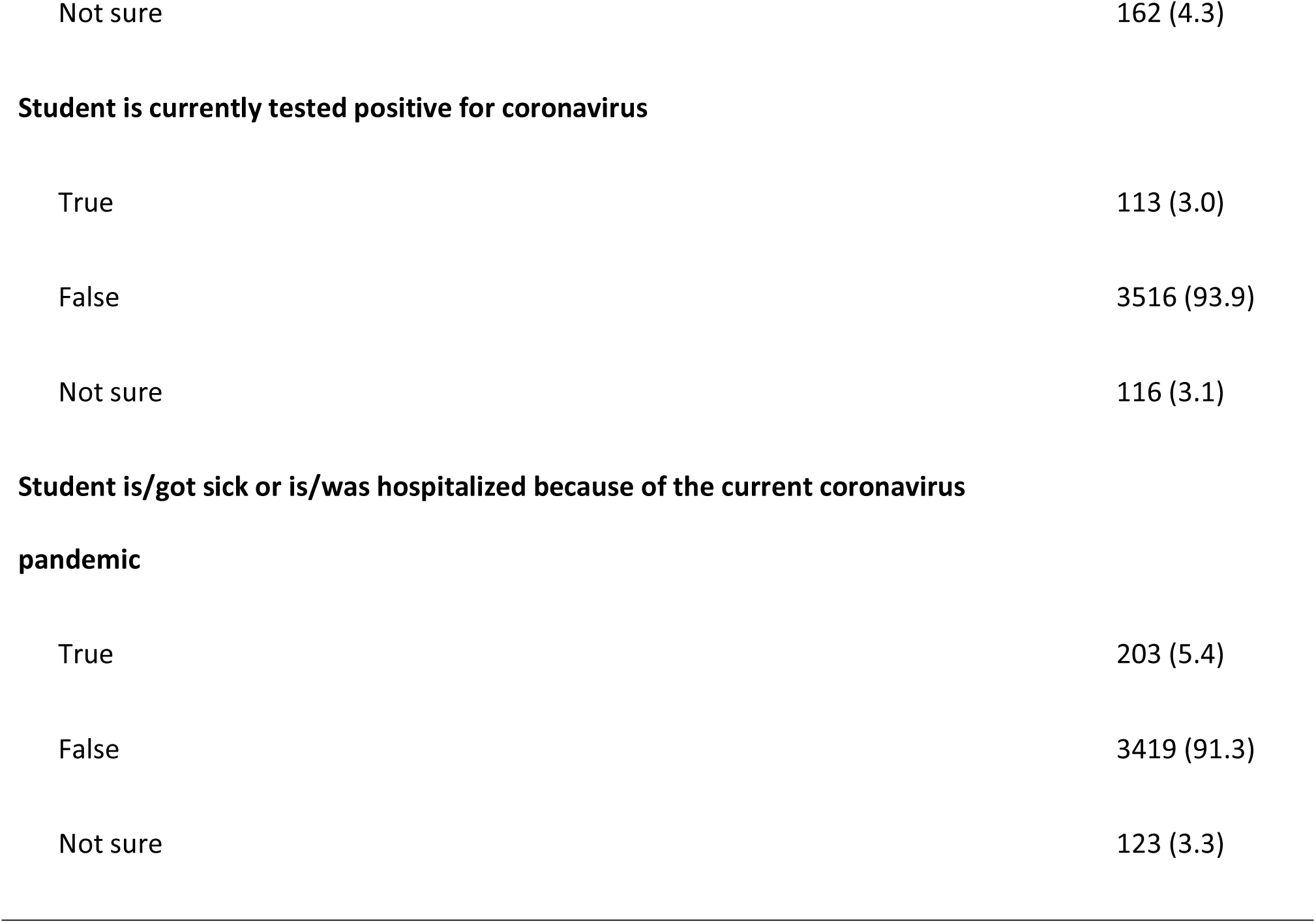
Epidemic related information for the sample (N=3,745)

| **Variables** | **Number (%)** |
| --- | --- |
| **A close family member has currently tested positive for coronavirus** |  |
| True | 532 (14.2) |
| False | 2998 (80.1) |
| Not sure | 215 (5.7) |
| **A close family member is/got sick or is/was hospitalized because of the current coronavirus infection** |  |
| True | 525 (14.0) |
| False | 3072 (82.0) |
| Not sure | 148 (4.0) |
| **You work around people who might have the current coronavirus** |  |
| True | 798 (21.3) |
| False | 2291 (61.2) |
| Not sure | 656 (17.5) |
| **A close family member died because of the current coronavirus** |  |
| True | 216 (5.8) |
| False | 3367 (89.9) |
| Not sure | 162 (4.3) |
| **Student is currently tested positive for coronavirus** |  |
| True | 113 (3.0) |
| False | 3516 (93.9) |
| Not sure | 116 (3.1) |
| **Student is/got sick or is/was hospitalized because of the current coronavirus pandemic** |  |
| True | 203 (5.4) |
| False | 3419 (91.3) |
| Not sure | 123 (3.3) |

The exploration of mental health symptoms indicated that the risk for PTSD was the most concerning issue (40.6%). Although anxiety and depression were found to be of lesser prevalence (23% and 17% respectively), they were still understood to be alarming.

Significant age and gender differences were found to be associated with mental health symptoms. A significantly higher proportion of participants who were late adolescents reported symptoms of two conditions (depression and anxiety) compared to participants in the younger age group (p<0.05). Additionally, a significantly higher proportion of female students reported symptoms of one condition-depression, anxiety, or risk for PTSD only-(p<0.05) or two conditions (p<0.05) (Table 3).

**Table 3:**
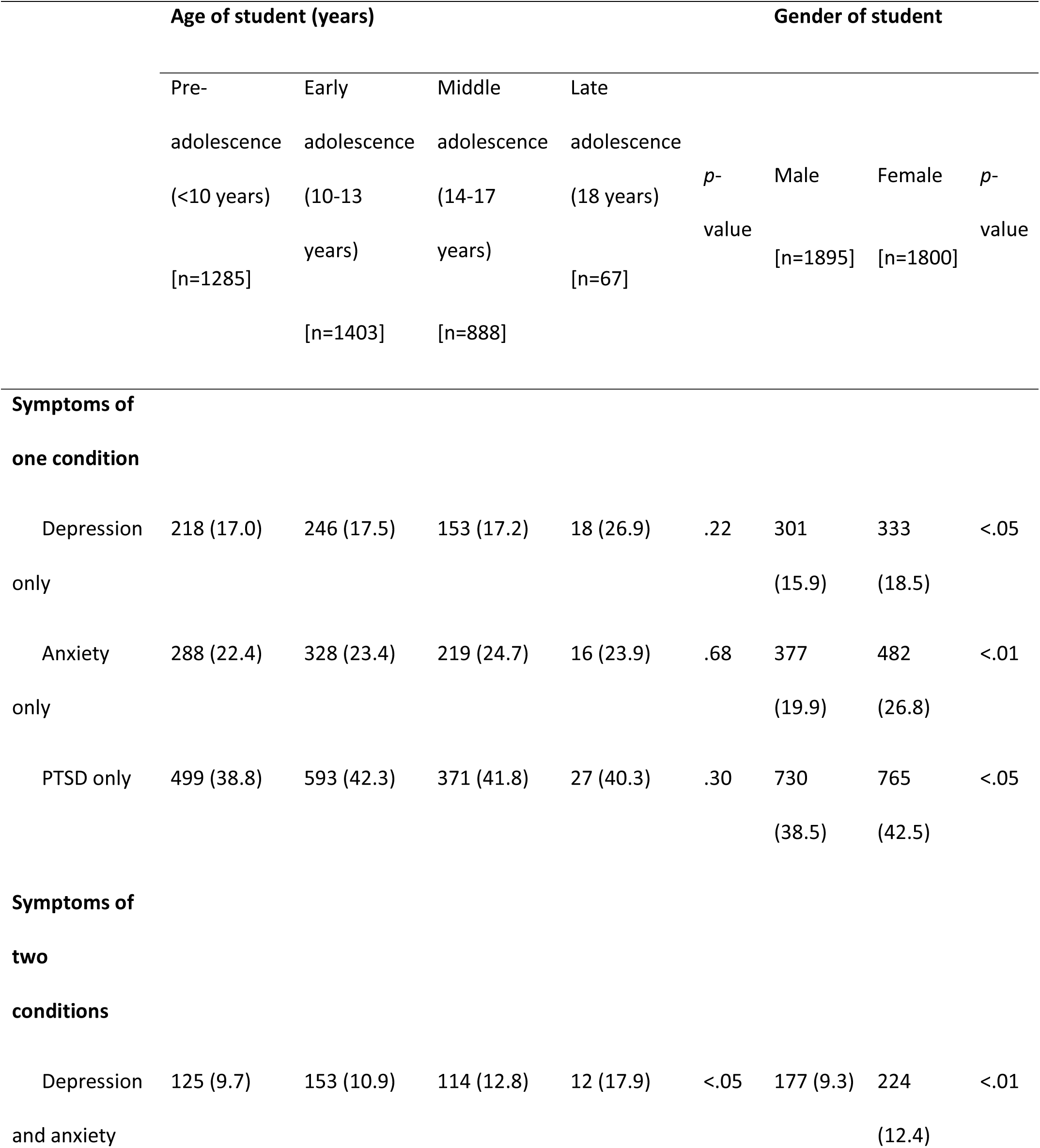
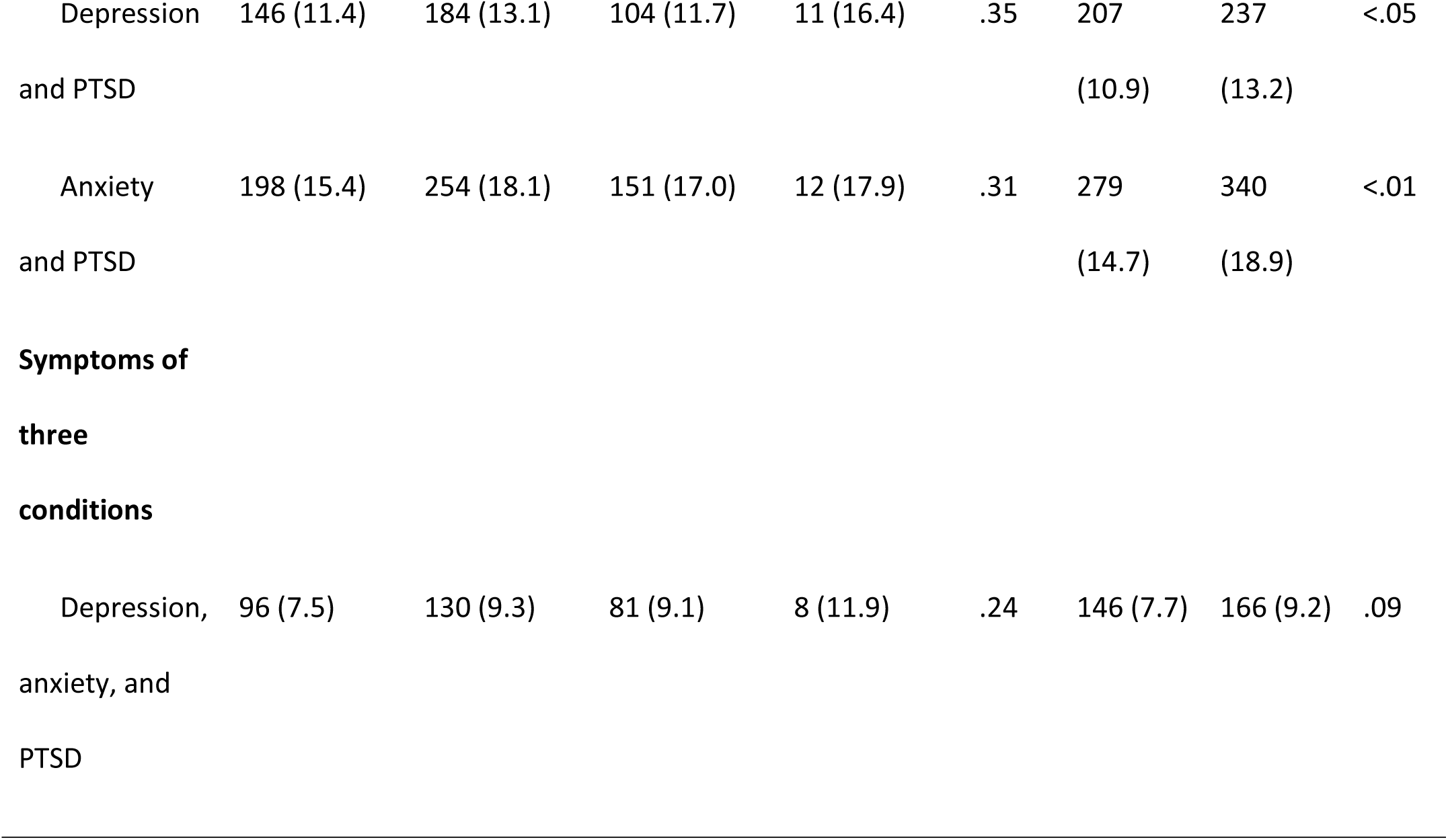
Distribution of symptoms by age and gender.

|  | Age of student (years) |  |  |  | Gender of student |  |  |  |
| --- | --- | --- | --- | --- | --- | --- | --- | --- |
|  | Pre-adolescence (<10 years) | Early adolescence (10-13 years) | Middle adolescence (14-17 years) | Late adolescence (18 years) | *p*-value | Male | Female | *p*-value |
|  | [n=1285] |  |  | [n=67] |  | [n=1895] | [n=1800] |  |
|  |  | [n=1403] | [n=888] |  |  |  |  |  |
| Symptoms of one condition |  |  |  |  |  |  |  |  |
| Depression only | 218 (17.0) | 246 (17.5) | 153 (17.2) | 18 (26.9) | .22 | 301 (15.9) | 333 (18.5) | <.05 |
| Anxiety only | 288 (22.4) | 328 (23.4) | 219 (24.7) | 16 (23.9) | .68 | 377 (19.9) | 482 (26.8) | <.01 |
| PTSD only | 499 (38.8) | 593 (42.3) | 371 (41.8) | 27 (40.3) | .30 | 730 (38.5) | 765 (42.5) | <.05 |
| Symptoms of two conditions |  |  |  |  |  |  |  |  |
| Depression and anxiety | 125 (9.7) | 153 (10.9) | 114 (12.8) | 12 (17.9) | <.05 | 177 (9.3) | 224 (12.4) | <.01 |
| Depression | 146 (11.4) | 184 (13.1) | 104 (11.7) | 11 (16.4) | .35 | 207 | 237 | <.05 |
| and PTSD |  |  |  |  |  | (10.9) | (13.2) |  |
| Anxiety | 198 (15.4) | 254 (18.1) | 151 (17.0) | 12 (17.9) | .31 | 279 | 340 | <.01 |
| and PTSD |  |  |  |  |  | (14.7) | (18.9) |  |
| **Symptoms of three conditions** |  |  |  |  |  |  |  |  |
| Depression, anxiety, and PTSD | 96 (7.5) | 130 (9.3) | 81 (9.1) | 8 (11.9) | .24 | 146 (7.7) | 166 (9.2) | .09 |

A more detailed examination of the relationship between pandemic related factors and mental health symptoms is shown in Table 4. Students who were reported to have medical problems were more likely to have mental health symptoms as compared to those with nil history (p<0.01). Those who reported a close family member was sick or hospitalized because of the coronavirus infection were more likely to report mental health symptoms as compared to those who reported the statement was false (p<0.05). In terms of the death of a close family member due to coronavirus infection, respondents who reported a close family member had died because of the coronavirus infection were more likely to show mental health symptoms as compared to those who did not (p<0.01).

**Table 4:**
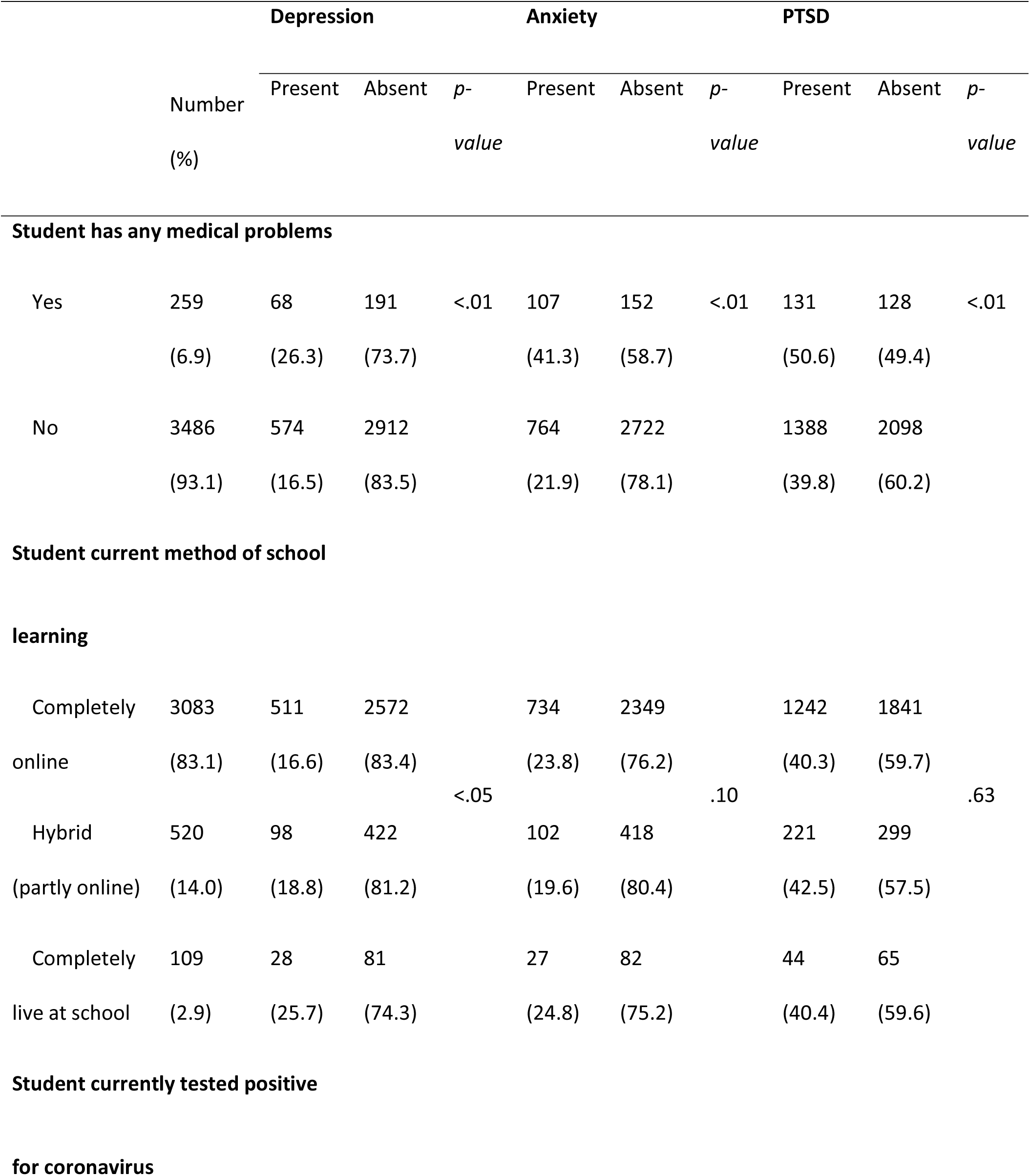
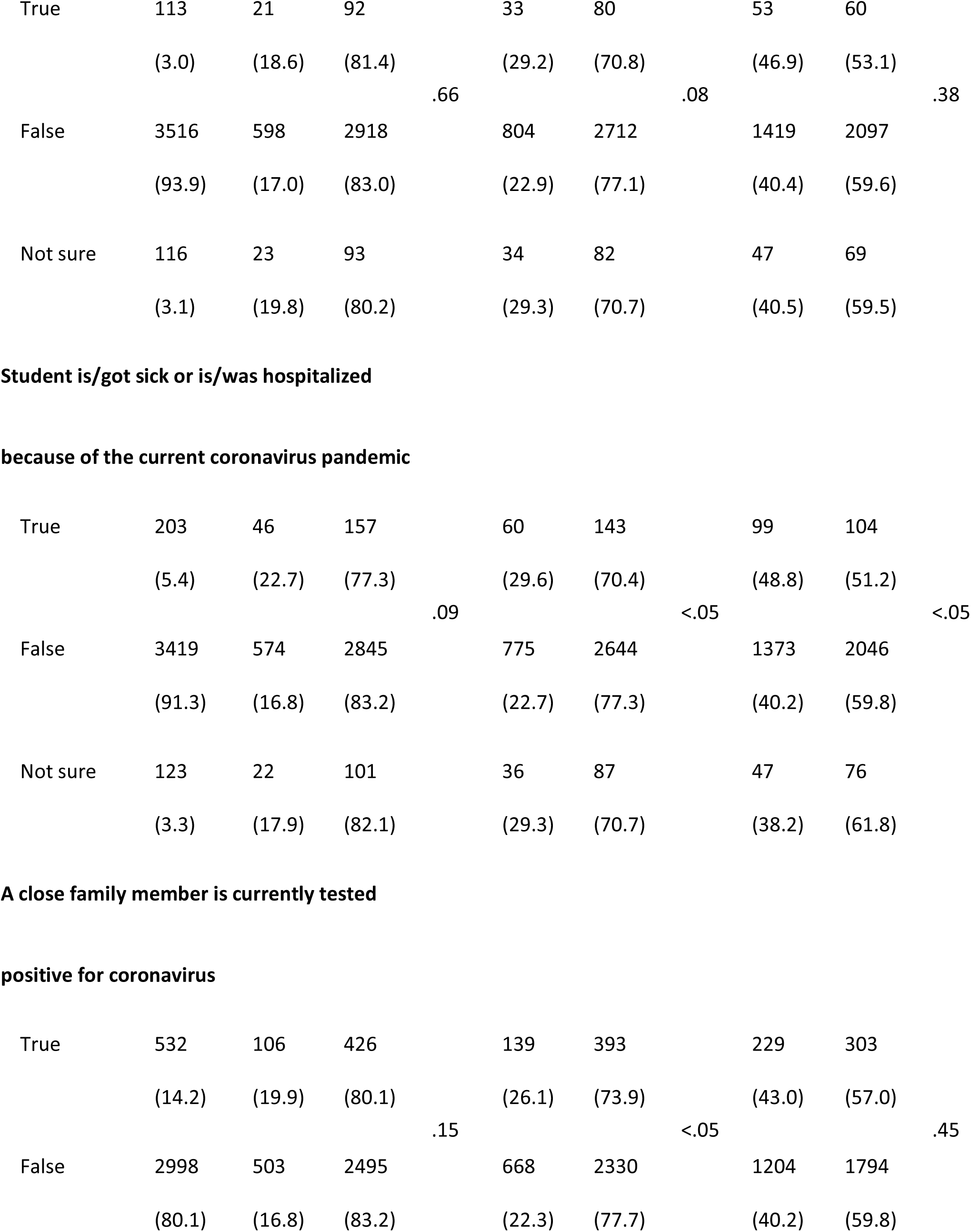
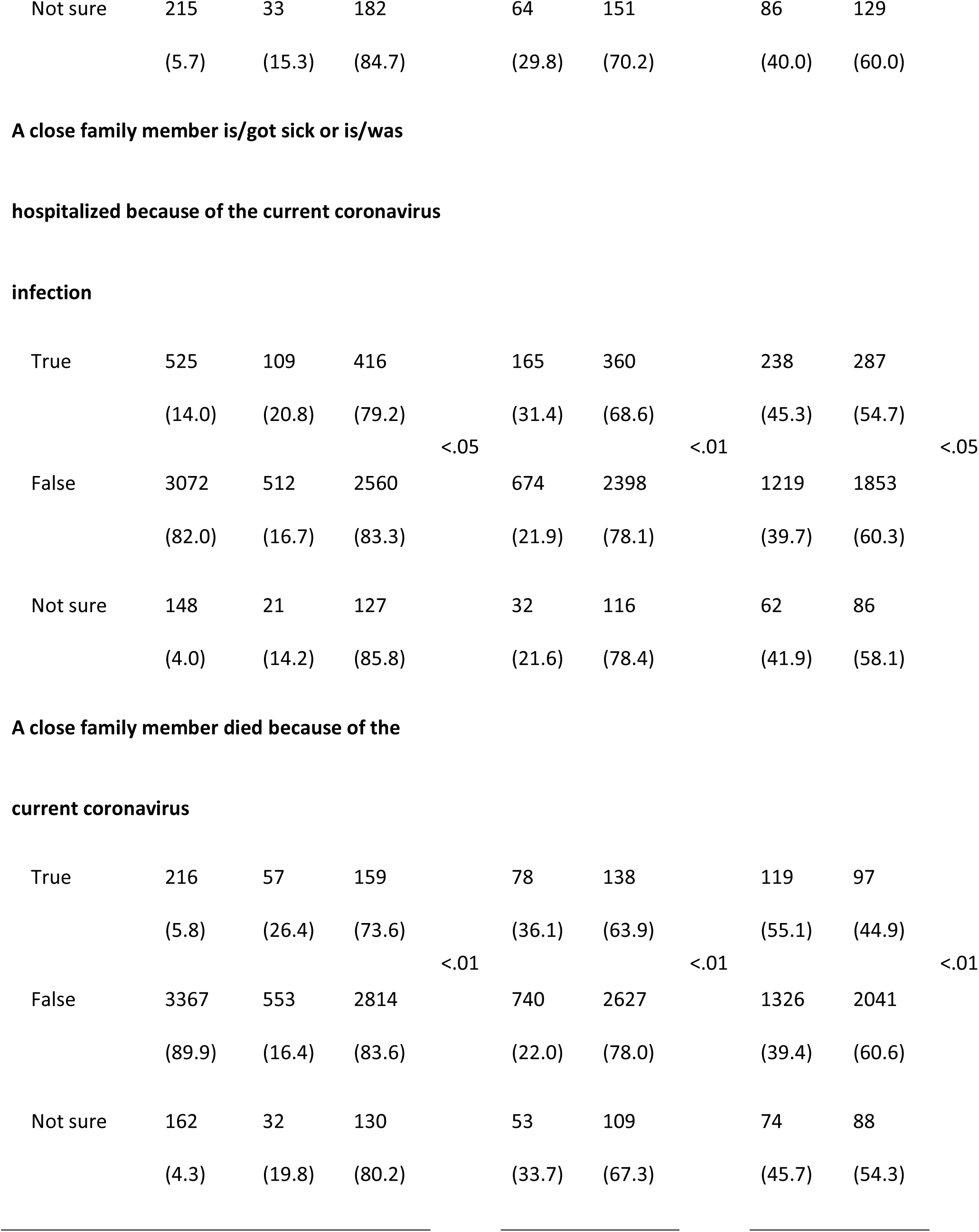
Prevalence of symptoms of depression, anxiety, PTSD stratified by epidemic-related factors.

Further regression analysis identified the factors associated with depression, anxiety, and risk for PTSD as shown in Table 5. Students who had any medical problems were 2.0 (95% CI 1.5-2.6) times more likely to display anxiety symptoms, and 1.3 (95% CI 1.0-1.8) times more at risk for PTSD. Likewise, participants who had experienced the death of a close family member due to coronavirus infection were 1.7 (95% CI 1.2-2.4) times more likely to report depression symptoms, 1.7 (95% CI 1.2-2.3) times more likely to have higher levels of anxiety, and 1.7 (95% CI 1.2-2.3) times more at risk for PTSD after adjusting for age, gender, and existing mental health-related problems.

**Table 5:**
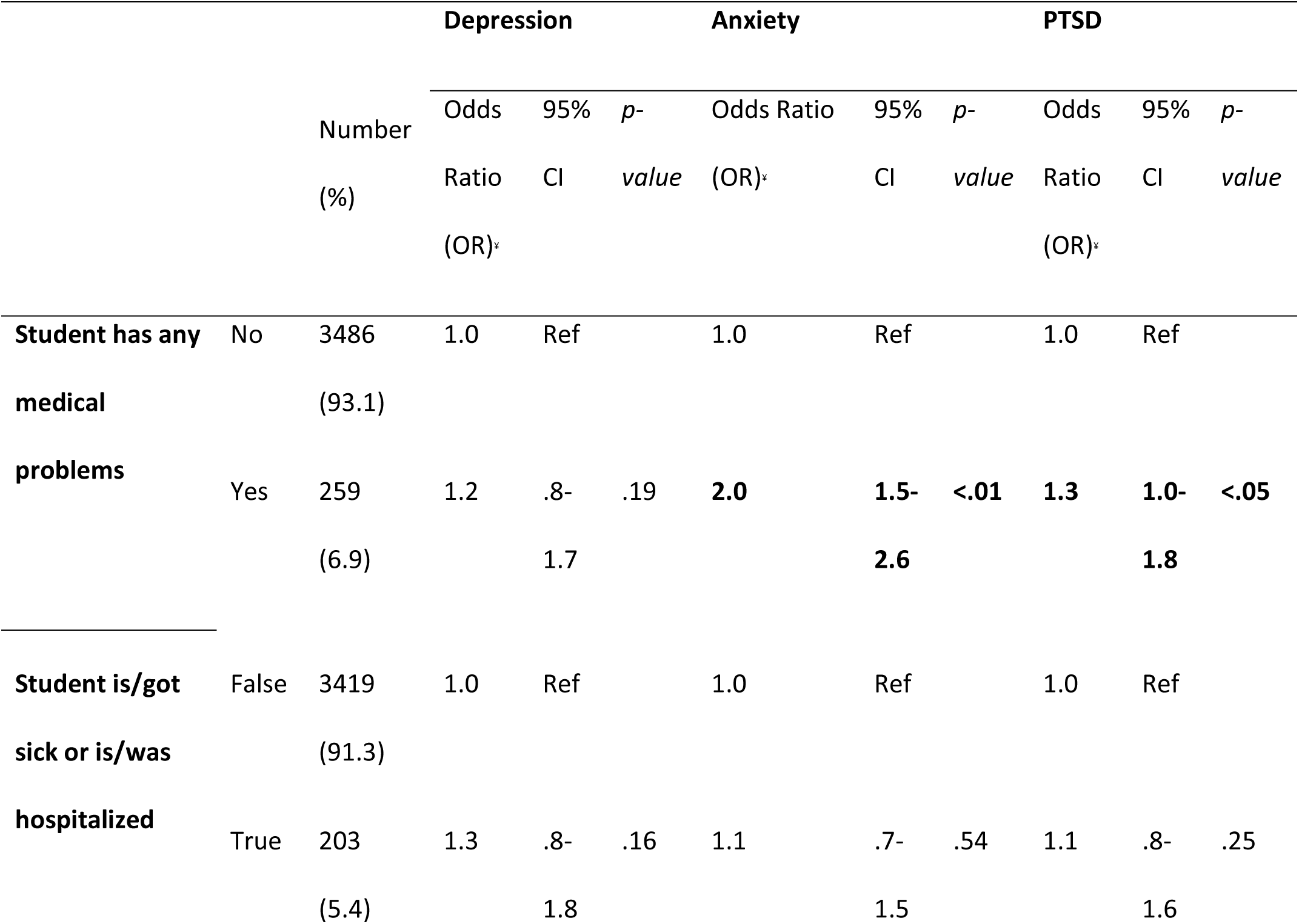
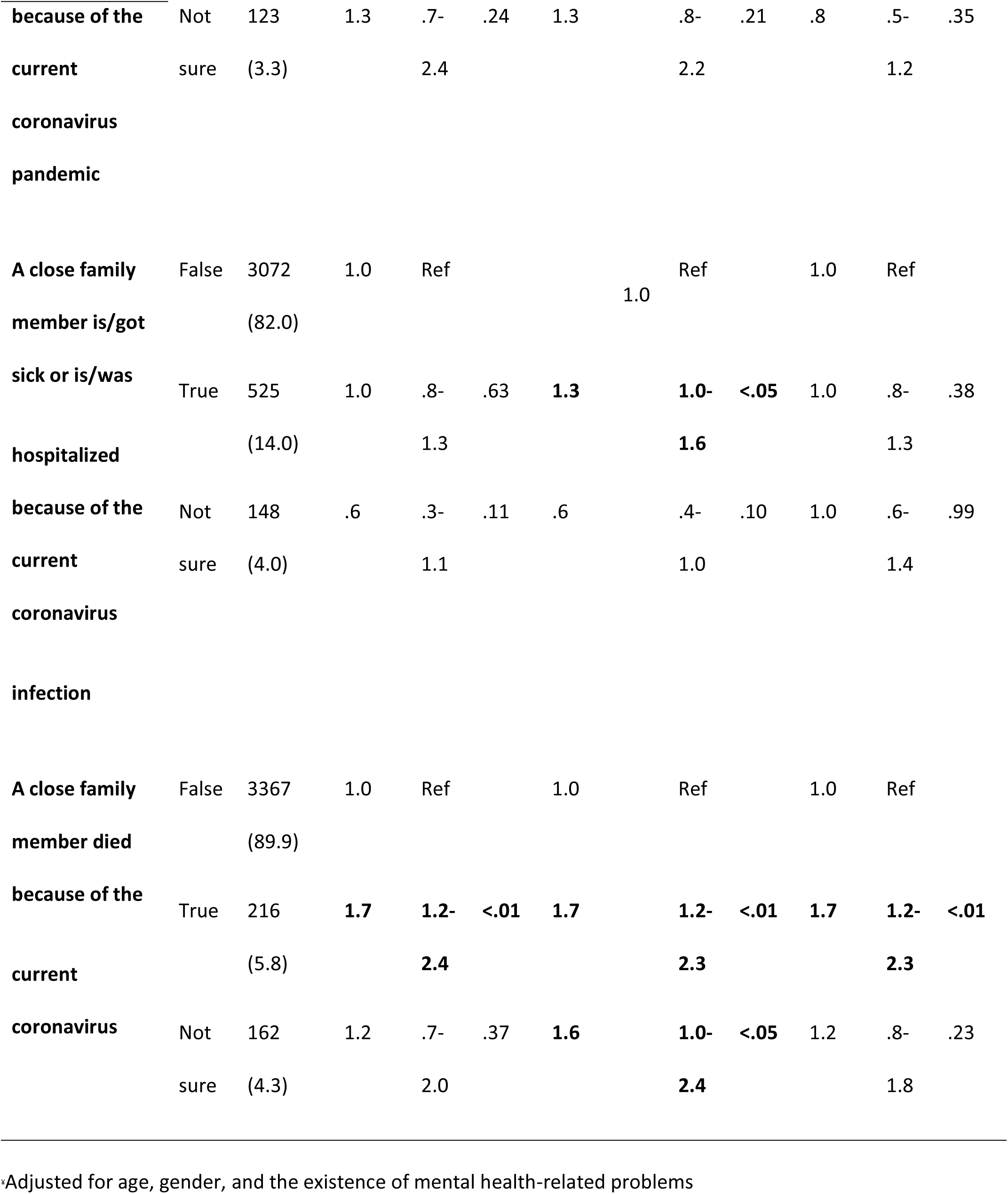
Predisposing factors to common mental health symptoms.

## Discussion

**T**his cross-sectional web-based study investigated the prevalence of mental health symptoms (depression, anxiety, and risk for PTSD among 3745 school students in the United Arab Emirates post lockdown of the COVID-19 pandemic. This study also identified the potential factors that predicted school students’ depression, anxiety, and risk for PTSD.

### Prevalence of anxiety, depression, and risk for PTSD

This study has adequate evidence that students in the UAE are exhibiting symptoms of mental health disorders in the post COVID-19 pandemic period and this is consistent with that of recent studies across the globe. In a comprehensive review of evidence, the WHO concluded that the world has experienced a 25% increase in the prevalence of anxiety and depression in the first year of the COVID-19 crisis. More precisely, the report revealed that the pandemic has particularly affected the mental health of young people who, more seriously, were found disproportionally at risk of suicidal and self-harming behaviors. Accordingly, the organization considered the COVID-19 pandemic a wake-up call to the world to set up mental health services [15-17]. Furthermore, a score of studies showed that adolescents were more under the weather during the pandemic compared to adults; these comparative results revealed that adolescents were more likely to report depression, anxiety, and PTSD [18, 19]. For the post-lockdown period, a study from China found out that PTSD symptoms were prevalent among a large-scale of adolescents in China (16.9%), followed by depression (12.8%), and anxiety (7.1%) [20].

### Gender and age differences

The results from this study highlight that female and older adolescents showed a significantly higher proportion of anxiety, depression, and risk for PTSD symptoms. These gender and age related differences are supported by a number of recent studies that showed female high school students at an increased risk of psychological stress [21] and at higher levels of anxiety and depressive symptoms during the pandemic [22]. Interestingly, a number of researchers have argued that a possible explanation for female high school students’ higher levels of anxiety, depression, and stress symptoms during the COVID-19 pandemic could be due to physiologic hormonal and bodily changes or a lack of coping mechanisms [23, 24].

### Past medical history and Family/social support

This study explored more associations between mental symptoms and existing medical conditions. Findings showed that students who had any medical problem were 2.0 (95% CI 1.5-2.6) times more likely to have anxiety, 1.3 (95% CI 1.0-1.8) times more likely to have PTSD. These results are supported by a recent post-lockdown study conducted in Germany which showed that children with complex chronic diseases were more likely to have mental health problems [25]. In another supportive study, researchers concluded that COVID-19-associated mental health risks were more likely to appear in children and adolescents with special needs [26]. Conventionally, social ties and family relationships are understood to have a significant impact on the maintenance of psychological well-being and reduce the risk for depression among adolescents [27-30]. The relevant findings from our study are supportive of this conviction; participants who reported that a close family member had tested positive for SARS-CoV-2, got ill, or was hospitalized due to the infection were more likely to have mental health disorders as compared to those who did not. Furthermore, participants who had experienced the death of a close family member due to coronavirus infection were 1.7 (95% CI 1.2-2.4) times more likely to have depression, 1.7 (95% CI 1.2-2.3) times more likely to have anxiety, and 1.7 (95% CI 1.2-2.3) times more likely to have PTSD.

In a nutshell, symptoms of anxiety, depression, and risk for PTSD were found in the UAE school students. Mental health services are needed to help those children cope and recover as the Covid-19 pandemic is receding. Special attention must be regarded to disadvantaged children and proactive plans should be put in place for potentially forthcoming threats.

### Strengths and Limitations

Healthcare entities in the UAE have been leveraging their resources to address robustly the mental health of children and adolescents even before the COVID-19 pandemic. However, the emergence of this global threat required nationwide collaboration to address the exigencies of the situation. Scientific mental health research in the UAE before the COVID-19 era was normally conducted at institutional level. As part of the national response to disaster management, mental health research in children and adolescents have been prioritized. A multi-disciplinary team from diverse government health regulators, the emergency and disaster management research team, healthcare service-providers (public and private sectors), and academia in the UAE collaborated to develop a solid ground of national data on the status of mental illness symptoms in the younger population. And what’s more, the data collection instrument was a mere innovative piece which garnered data on eight different scales (sociodemographic, COVID-19, prosocial behavior-SDQ, psychosocial Impact-PIS, mood and feelings-MFQ, child anxiety-SCARED, impact of events-CRIES, loneliness-UCLA-LS). A pilot test was run to ensure the validity and reliability of the hybrid tool and adjustments were made accordingly. The strong collaboration that EHS steered led to cultivating the inputs from parents and school students from the seven emirates (N=3762). In addition, the large sample size maximized the statistical significance, as well as minimized the statistic’s margin of error. Eventually, robust data sets were generated which would serve multiple disciplines connected to children and adolescents’ health (physical and mental health, educational, social, and economical). One more proven strength of this study comes from the literature review which revealed that such work is the first of its kind in the UAE and its findings shall serve as scientific evidence on the magnitude and nature of work needed in children and adolescents’ mental health. Conversely, the main limitation of this work comes from the extraordinary abundance of data variables which made it impossible for researchers to incorporate them all in a single study. This shortcoming can be rectified by calling upon researchers to derive from this data bank a series of other relevant studies that would inform the community of mental health professionals, decision makers, and policy developers in their intent to promote the well-being and overall mental health of children and adolescents. A second logistic limitation comes from the limited pre-pandemic data on the status of children and adolescents mental health in the UAE which gave little chance to explore a causal relationship. Finally, the data was collected using self-reported measures without any clinical encounters.

## Conclusion

Lockdown, social restrictions, school closures, fear, sickness, death, and all the COVID-19 pandemic mood have placed significant mental health pressures on school students and their families across worldwide. Our findings highlight the prevalent mental symptoms faced by school students in the UAE post lockdown of the current COVID-19 pandemic. Moreover, the findings extend the current understanding of the underlying factors that contribute to the development of anxiety, depression, and PTSD among school students. The current study primarily highlights the particular importance of identifying school students who are at high risk of mental health disorders. Likewise, it provides information for family, school administrators, and policymakers to work jointly for a sustainable and socially supportive environment that maintains and promotes the well-being and overall mental health of school students with particular considerations to emergencies and disasters preparedness.

The results from this study provide significant guidance for the development of psychological support strategies for school students. Particular attention should be paid to groups at higher risks (late adolescents, female students, and those living within disturbed family relationships and suffering from complex chronic diseases). In addition, students who suffered a traumatic experience during the pandemic such as becoming critically ill, losing a loved one, or being hospitalized over an extended period of time should receive additional attention. There is a need to continuously evaluate the mental wellness of students, to take specific measures to help students cope with the evolving learning environment, to identify the psychological needs of students instantly and at an individual level, to provide psychological interventions for students suffering from psychological distress, to train teachers to understand the verbal and non-verbal cues of students that tell about potential mental sufferings, and to solidify communications between school and family. On a broader spectrum, this study calls upon healthcare and non-healthcare entities, policy-makers, and the civil society that invest in shaping the future to develop a comprehensive school mental health program that promotes the well-being of youth and strengthens the resilience of these minors in the face of the forthcoming uncertainties and threats.

## Data Availability

All data produced in the present study are available upon reasonable request to the authors

## Acknowledgements

We thank Dr. Aji Gopakumar, Dr. Mariam Jaafar, and Ms. Amna AlDhmanie from the Data and Statistics Department –Emirates Health Services Establishment for assisting in publishing this research. Likewise, we acknowledge Ms. Ahlam Al Maskari for assisting in data collection at Abu Dhabi Public Health Center and the research team at the Statistics and Research Center-Ministry of Health and Prevention.

